# Some markers of placental inflammation in pregnant women with gestational diabetes

**DOI:** 10.1101/2024.10.11.24315095

**Authors:** Thiago Paes de Barros De Luccia

## Abstract

**Background:** analysis of fetal/placental components of women with gestational diabetes presented a slightly inflammatory profile, compared with non-diabetic pregnant women in previous studies by our group. Leptin, resistin and IL-6 are involved in the inflammatory process while adiponectin can act in anti-inflammatory processes.

**Purpose:** Since both obesity and gestational diabetes are associated with inflammatory conditions, through these mediators we seek to evaluate systemic patterns in pregnant women and fetal patterns of this possible inflammation.

**Materials and Methods:** Three adipokines (leptin, resistin and adiponectin) and one cytokine (IL-6) were studied in three different compartments (maternal serum, fetal serum and amniotic membrane culture supernatant). Four pregnant groups were analyzed (eutrophic, overweight, obese and gestational diabetics (GDM)). Maternal and fetal serum and amnion membrane biopsies were collected from 20 GDM and 28 normoglycemic subjects (Controls) who underwent elective cesarean sections.

**Results:** We found a higher production of IL-6 in the culture supernatant of the amniotic membrane of obese pregnant women and more significantly in pregnant women with gestational diabetes. We did not observe correlation between the levels of mediators detected in the three compartments (mother serum, fetal serum and culture supernatant of amniotic membrane).

**Conclusion:** Mainly in the amniotic membrane of pregnant women with GDM, a slight increase in inflammatory markers was observed.

## Introduction

In previous studies carried out by our group, analysis of fetal/placental components of women with gestational diabetes presented a slightly inflammatory profile, compared with non-diabetic pregnant women^1,2^. In this article we will present supplementary data obtained after the aforementioned publications. In this new data, we added a more detailed analysis of the behavior of markers (leptin, adiponectin, resistin and IL-6) according to the pregnant women’s BMI. Since both obesity and gestational diabetes are associated with inflammatory conditions, through these mediators we seek to evaluate systemic patterns in pregnant women and fetal patterns of this possible inflammation.

## Methods

Three adipokines (leptin, resistin and adiponectin) and one cytokine (IL-6) were studied in three different compartments (maternal serum, fetal serum and amniotic membrane culture supernatant). Four pregnant groups were analyzed (eutrophic, overweight, obese and gestational diabetics). Maternal and fetal serum and amnion membrane biopsies were collected from 20 GDM and 28 normoglycemic subjects (control) who underwent elective cesarean sections during the years 2017 to 2020. The amnion membrane was manually peeled from the fetal membranes and placed in a 12-well culture for 24 hours at 37°C in a 5% CO_2_ atmosphere and 95% air humidity (details about amniotic membrane culture in Menon et al., 2011^3^). The cytokine and adipokines were evaluated in serum and amnion culture supernatant samples (Luminex and ELISA). The detailed methodology of the work can be consulted in a previously published article^1^. The participants were recruited at the Obstetric Center of Hospital São Paulo-Brazil and of the Hospital do Servidor Público Municipal do Estado de São Paulo-Brazil (Ethics and Research Committee of UNIFESP CAAE 80728417.8.0000.5505).

## Results

Here we will only present the most consistent results, as in some compartments (MS, FS and CS) measuring all markers was not possible. IL-6 measurements, for example, were better in the fetal compartment. We were unable to achieve good results in detecting this cytokine in maternal and fetal serum. Figure 1 shows these results of this cytokine in amniotic membrane culture. In the graph (**Figure 1**) we see a greater production of IL-6 in the membrane of obese pregnant women and those with gestational diabetes.

**Figure 1-.**
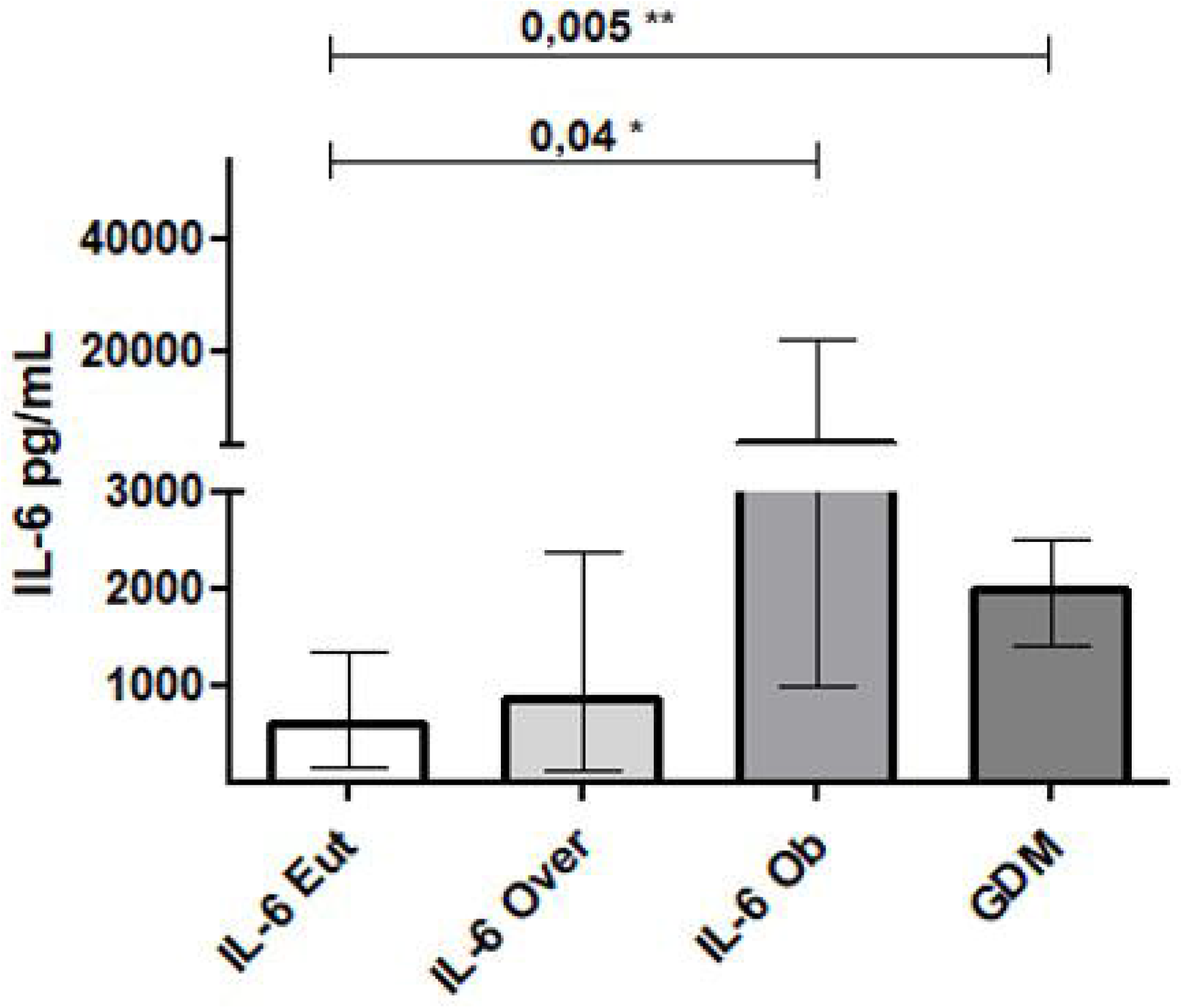
IL-6 (culture supernatant - CS). Eut=eutrophic, Over=overweight, Ob=obese and GDM=gestational diabetics.

In table 1 we have the most consistent results in all groups of pregnant women. In **Figure 2-A** and **Figure 2-B** we present the leptin/adiponectin ratio in pregnant controls and pregnant women with gestational diabetes.

**Table 1-.** Adipokines and cytokines: median (interquartile range) - Mann Whitney test. BMI: mean (standard deviation) – t test. * significant difference. GDM: gestational diabetes *mellitus*. MS: mother serum; FS: fetal serum; CS: culture supernatant of amniotic membrane.

| Variable | Eutrophic (14) | Overweight (8) | Obese (6) | GDM (20) | Eut x<br>Over | Eut x Ob | Eut x<br>GDM | Over x<br>Ob | Over x<br>GDM | Ob x<br>GDM |
| --- | --- | --- | --- | --- | --- | --- | --- | --- | --- | --- |
| mean BMI<br>(kg/m<sup>2</sup>) | 22,3 (2,2) | 26,3 (0,9) | 32,8 (3,6) | 28,4 (5,1) | **<0.0001\*** | **<0.0001\*** | **<0.0001\*** | **<0.0001\*** | 0,0780 | **0,0091\*** |
| Adiponectin MS<br>(µg/mL) | 3,7 (1,9-4,2) | 2,2 (1,2-3,3) | 1,1 (1,0-1,5) | 2,8 (1,2-4,2) | 0,1051 | **0,0023\*** | 0,6430 | **0,0281\*** | 0,3493 | **0,0061\*** |
| Adiponectin FS<br>(µg/mL) | 3,4 (1,1-6,4) | 5,0 (1,7-6,5) | 1,1 (0,9-2,1) | 8,1 (3,5-14,5) | 0,5832 | 0,0564 | **0,0090\*** | **0,0152\*** | **0,0491\*** | **0,0007\*** |
| Leptin MS<br>(ng/mL) | 0,2 (0,1-0,6) | 40,4 (11,1-63,1) | 89,1 (65,3-182,5) | 39,7 (19,5-56,2) | **0,0003\*** | **0,0003\*** | **<0.0001\*** | **0,0041\*** | 1,0000 | **0,0010\*** |
| Leptin FS<br>(ng/mL) | 6,1 (4,0-17,0) | 12,5 (5,5-18,5) | 16,6 (13,3-29,8) | 17,5 (7,8-30,4) | 0,3040 | **0,0324\*** | **0,0261\*** | 0,1605 | 0,3423 | 0,8365 |
| Leptin CS<br>(ng/mL) | 0,25 (0,1-0,6) | 0,44 (0,08-1,4) | 0,46 (0,2-0,7) | 0,41 (0,1-0,7) | 0,9109 | 0,4333 | 0,6132 | 0,7308 | 0,9702 | 0,5919 |
| Resistin MS<br>(ng/mL) | 9,1 (6,3-12,3) | 8,6 (8,0-12,1) | 16,6 (14,1-17,7) | 6,6 (4,3-10,5) | 0,8255 | **0,0044\*** | 0,2048 | **0,0256\*** | 0,0579 | **0,0036\*** |
| Resistin FS<br>(ng/mL) | 11,6 (9,3-18,4) | 19,6 (11,7-27,0) | 17,2 (14,8-21,9) | 13,4 (12,0-15,3) | 0,1148 | 0,0872 | 0,9078 | 1,0000 | 0,2573 | **0,0307\*** |
| Resistin CS<br>(ng/mL) | 1,3 (0,7-2,1) | 0,5 (0,2-1,8) | 0,8 (0,7-1,0) | 1,6 (0,9-2,1) | 0,0998 | 0,2055 | 0,8178 | 0,5237 | 0,1803 | 0,0611 |
| IL-6 CS (pg/mL) | 612,8 (153-1364) | 881,0 (136-2381) | 3424,0 (999-22071) | 2016,0 (1416-2515) | 0,2423 | **0,0426\*** | **0,0053\*** | 0,2343 | 0,3595 | 0,6128 |

**Figure 2 - A.**
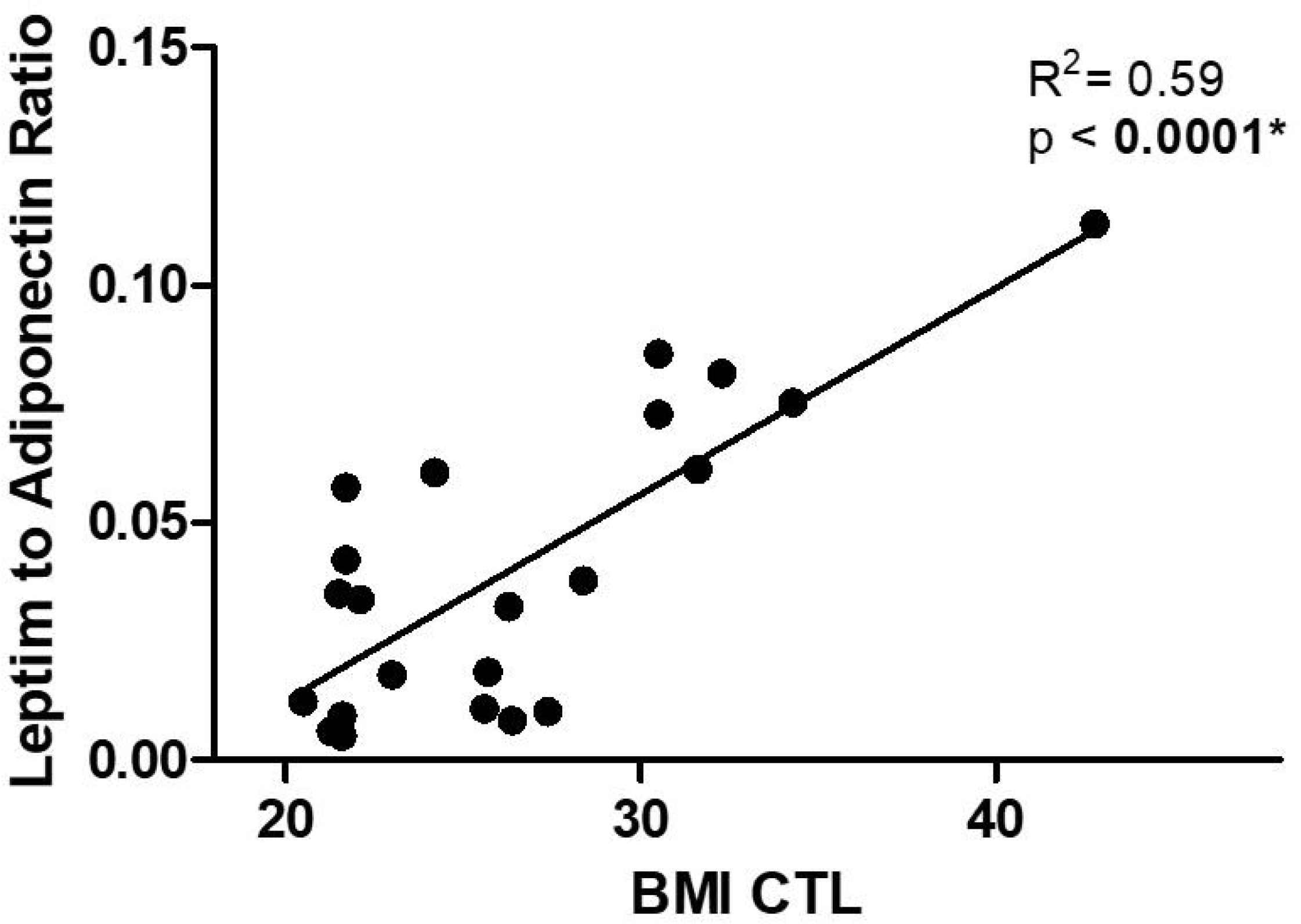
correlation between leptin to adiponectin ratio and BMI in Controls.

**Figure 2 - B.**
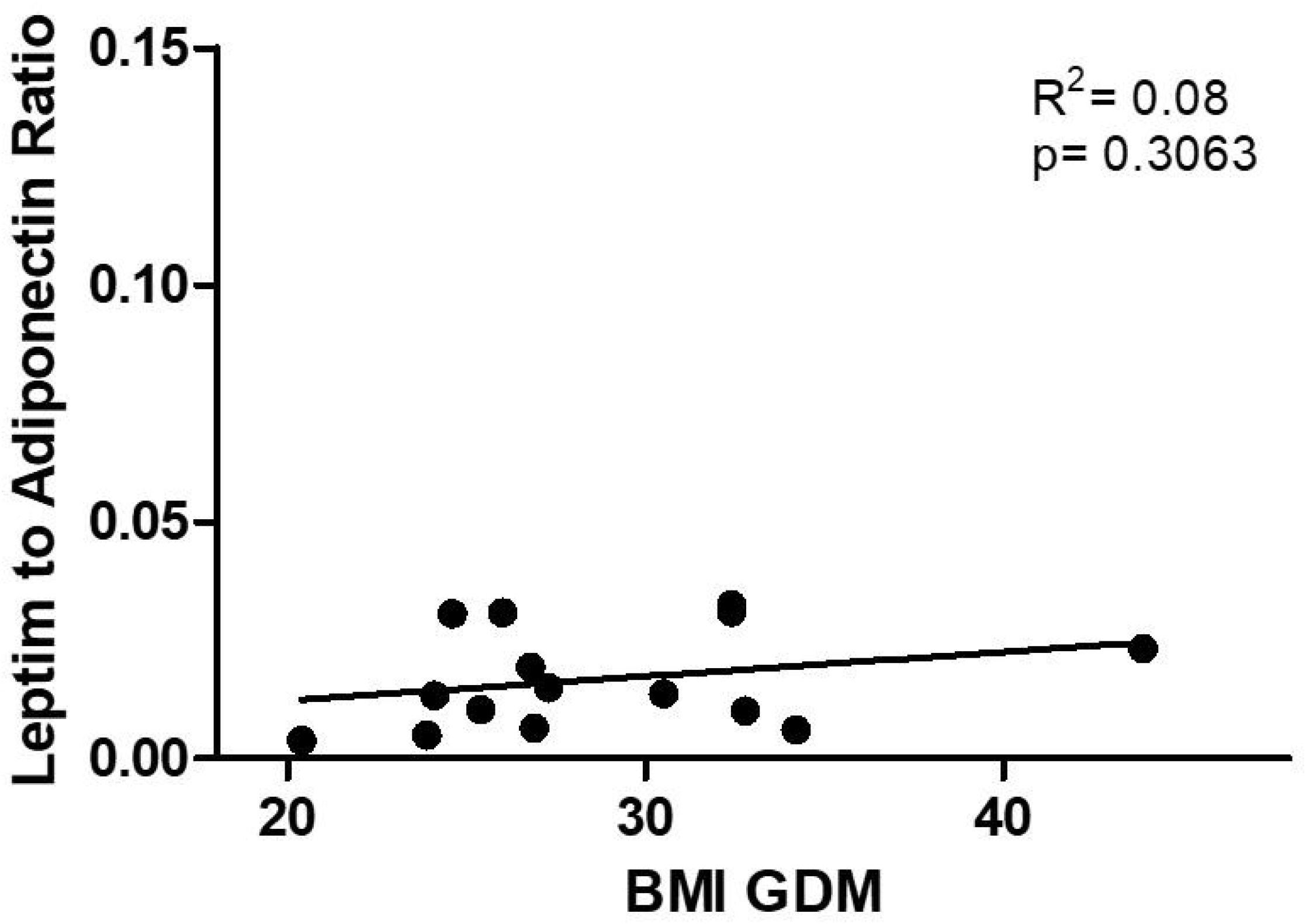
correlation between leptin to adiponectin ratio and BMI in GDM.

## Discussion

Ayina et al. showed that leptin to adiponectin ratio was significantly and positively correlated with the body mass index (r = 0.669, p < 0.0001)^4^. Higher levels of this ratio are associated with greater cardiovascular risks ^5-7^. In our study, in the GDM group, the positive correlation did not happen, maybe because the variation of BMI was smaller in this group. In the GDM group, the BMI was more similar to the overweight BMI group.

In relation to adipokines, there was a certain similarity in dosages between overweight pregnant women and gestational diabetics, suggesting that BMI variation is an important factor controlling adipokines. If we look at table 1, we see that there was no significant difference in dosages between the GDM group and overweight controls. More studies with a larger number of participants need to be carried out to understand the role of gestational diabetes in changing these markers (with BMI matching).

In studies on intra-amniotic inflammation, IL-6 was shown to be a good marker of inflammation^8^. The increase in this marker in amniotic fluid and cervical fluid has also been associated with preterm birth^9^. In this scenario, the amniotic membrane may play a role in the production of this cytokine. Our data showed greater amounts of IL-6 produced by the amniotic membrane in obese women and more significantly in gestational diabetics. However, there is no clear association between obesity/gestational diabetes and premature birth, but rather with advanced maternal age and premature birth^10^.

In our previous studies, we did not observe correlation between the levels of mediators detected in the three compartments (mother serum, fetal serum and culture supernatant of amniotic membrane). Mainly in the amniotic membrane of pregnant women with GDM, a slight increase in inflammatory markers was observed.

## Data Availability

All data produced in the present study are available upon reasonable request to the authors

https://pubmed.ncbi.nlm.nih.gov/33836321/

## Notes

### Competing Interest Statement

The authors have declared no competing interest.

### Clinical Protocols

https://pubmed.ncbi.nlm.nih.gov/33836321/

### Funding Statement

This work was supported by the Sao Paulo Research Foundation (FAPESP) [Grant number 2016/16807-9]; Conselho Nacional de Desenvolvimento Cientifico e Tecnologico (CNPq) [Grant number 302969/2019-5]; and by the Coordenacao de Aperfeicoamento de Pessoal de Nivel Superior - Brasil (CAPES) - Finance Code 001.

### Author Declarations

Obstetric Center of Hospital Sao Paulo-Brazil. Ethics and Research Committee of UNIFESP CAAE 80728417.8.0000.5505

